# Effects of midfoot joint mobilization on perceived ankle-foot function in chronic ankle instability. A crossover clinical trial

**DOI:** 10.1101/2021.12.08.21267445

**Authors:** Abbis Jaffri, John J. Fraser, Rachel M. Koldenhoven, Jay Hertel

## Abstract

**Background:** Chronic ankle instability (CAI) is a complex clinical entity that commonly includes ankle-foot impairment.

**Objective:** To investigate the effects of midfoot joint mobilizations and a one-week home exercise program (HEP) compared to a sham intervention and HEP on pain, patient-reported outcomes (PROs), ankle-foot joint mobility, and neuromotor function in young adults with CAI.

**Methods:** Twenty participants with CAI were instructed in a stretching, strengthening, and balance HEP and were randomized *a priori* to receive midfoot joint mobilizations (forefoot supination, cuboid glide and plantar 1^st^ tarsometatarsal) or a sham laying-of-hands. Changes in foot morphology, joint mobility, strength, dynamic balance, and PROs assessing pain, physical, and psychological function were assessed pre-to-post treatment and one-week following. Participants crossed-over to receive the alternate treatment and were assessed pre-to-post treatment and one-week following. Linear modelling was used to assess changes in outcomes.

**Results:** Participants who received midfoot mobilization demonstrated significantly greater perceived improvement immediately posttreatment in the single assessment numeric evaluation (Sham: 5.0±10.2%; Mobilization: 43.9±26.2%; β: 6.8 p<0.001, Adj R^2^:0.17) and Global Rating of Change (Sham: −0.1±1.1; Mobilization: 1.1±3.0; β: 1.8 p=0.01, Adj R^2^:0.12). Following the mobilization intervention, participants demonstrated greater improved rearfoot inversion mobility (Sham: 4.4±8.4°; Mobilization: −1.6±6.1°; β: −6.37, p=0.01, Adj R^2^:0.19), plantarflexion mobility (Sham: 2.7°±6.4; Mobilization: −1.7°±4.3; β: −4.36, p=0.02, Adj R^2^:0.07), and posteromedial dynamic balance (Sham: 2.4±5.9%; Mobilization: 6.0±5.4%; β: 3.88, p=0.04, Adj R^2^:0.10) compared to controls at 1-week post-treatment.

**Conclusion:** Participants with CAI who received midfoot joint mobilization had greater perceived improvement and physical signs that may benefit this clinical population.

## INTRODUCTION

Ankle sprains are the most common injury in orthopaedics and sport medicine.^1^ Individuals who incur ankle sprains often have difficulty returning to pre-injury functional levels and frequently experience injury recurrence.^2^ While many recover without residual deficit, 40% of the people who have an ankle sprain progress to develop chronic ankle instability (CAI) at least 12 months beyond the index ankle sprain.^3^ CAI is a clinical condition that includes mechanical and functional instability, along with residual symptoms in the ankle after a lateral ankle sprain.^4^ Mechanical deficits may include joint laxity, arthrokinematics restrictions, osteoarthritic changes, and synovial changes.^2,5^ Functional insufficiencies constitute proprioceptive deficits, loss in muscle strength, and neuromuscular alterations.^2,5^

Foot impairments such as altered joint motion, ligamentous laxity and pain, and strength deficits contribute to activity limitation and diminished quality of life in patients with CAI.^6^ Moreover, midfoot injury has been reported in patients who suffered lateral ankle sprains with 41% having midfoot ligamentous involvement and 33% having midfoot capsular injury.^7^ Manual Therapy (MT) is often used by rehabilitation specialists following injury to improve symptoms, range of motion, and sensorimotor function as a complementary treatment during return to activity.^8^ Additionally, the cascade of temporal neurophysiological effects fostered by joint mobilization may have specific beneficial benefits during treatment in this clinical population.^9^ As such, manual therapy interventions to include joint mobilization are recommended for inclusion in care when coupled with supervised or unsupervised exercise,^10^ While there is evidence for the use MT techniques in this population, study focus has been primarily on the treatment of the talocrural joint with the foot mostly ignored.^9^

Since the rates of CAI do not appear to be decreasing and the persistent perception of “it’s just an ankle sprain,” it is important to explore complementary treatments that may provide medicinal mechanical, neurophysiological, and psychological effects.^2^ In a related study of individuals with post-acute lateral ankle sprains, midfoot mobilizations was found to have positive psychological effects in regard to symptoms and function, facets that have are important in the mediation of intrinsic factors that are important in pain, treatment compliance, and functional outcomes.^11^ Therefore, the purpose of this crossover clinical trial was to investigate the effects of midfoot joint mobilization and a one-week home exercise program (HEP) compared to a sham intervention and HEP on patient-reported and clinical measures in young adults with CAI.

## METHODS

### DESIGN

This study is the second crossover clinical trial in a research line assessing the effects of midfoot mobilization in individuals with a history of ankle sprain, with the methods previously reported.^11^ A laboratory-based, crossover clinical trial was performed where the independent variable was treatment (50% allocated to initially receive joint mobilization, 50% allocated to initially receive sham). The primary dependent variables were changes in patient-reported pain and function, foot morphology (foot mobility magnitude, arch height flexibility), joint motion (weight-bearing dorsiflexion, rearfoot goniometry, forefoot inclinometry, 1^st^ metatarsal displacement), strength (handheld dynamometry), and dynamic balance (Star Excursion Balance Test, SEBT) immediately post-treatment and one-week following. Crossover design was selected over a parallel randomized control trial to ensure the individual factors of joint phenotype, injury heterogeneity, and psychological factors were accounted for in the design.^11^ The trial was registered with the National Institutes of Health (NCT02697461). The study was approved by the Health Science Research Institutional Review Board at the University of Virginia.

### PARTICIPANTS

Participants in this study were part of a larger research effort assessing foot impairment in individuals with and without CAI.^8,9,11^ A convenience sample of 20 recreationally-active individuals (9 males, 11 females) aged 18-35 with a recent history of LAS were recruited at a public university. Recreationally active was defined as participation in some form of physical activity for at least 20-minutes per day, at least three times a week. Participants must have incurred an ankle sprain ≥ 12-months prior to the study, experienced perceived or episodic giving way and reported deficit on the Identification of Functional Ankle Instability^5^ (IdFAI>10), Foot and Ankle Ability Measure (FAAM) ADL <90 and FAAM-Sport < 85.^14^ Participant demographics and self-report measures are in **Table 1**. Individuals were excluded if they had an ankle sprain within 8 weeks prior to the study, a self-reported history of leg or foot fracture, neurological or vestibular impairment that affected balance, diabetes mellitus, lumbosacral radiculopathy, soft tissue disorders such as Marfan syndrome or Ehlers-Danlos syndrome, any absolute contraindication to manual therapy, or if they were pregnant. Participants who met inclusion criteria provided informed consent. **Figure 1** details the CONSORT^23^ flow chart from recruitment to analysis.

**Table 1.** Demographic, injury history, and patient-reported outcome measures in individuals with chronic ankle instability

|  | Participants<br>allocated to receive<br>sham intervention<br>first (n=11)<br>4 males 7 females | Participants allocated<br>to receive midfoot<br>mobilization first<br>(n=9)<br>4 males 5 females |  |
| --- | --- | --- | --- |
| | Mean ± SD | | *p* |
| Age (years) | 21.3±6.3 | 20.3±1.3 | .64 |
| Height (cm) | 163.8±6.9 | 171.7±10.4 | .07 |
| Weight (kg) | 70.5±13.6 | 70.3±16.1 | .98 |
| BMI (kg/m<sup>2</sup>) | 26.3±4.8 | 23.7±4.0 | .21 |
| Foot Posture Index | 4.6±4.2 | 4.2±2.9 | .84 |
| Ankle sprains (n) | 6.0±7.5 | 4.4±2.1 | .52 |
| IdFAI | 23.8±3.6 | 23.3±3.5 | .77 |
| Kinesiophobia (TSK-11) | 23.7±5.6 | 21.2±6.6 | .38 |
| cm=centimeters, kg=kilograms, BMI=body mass index, IdFAI=Identification of Functional Ankle Instability, TSK=Tampa Scale Kinesiophobia |  |  |  |

**Figure 1.**
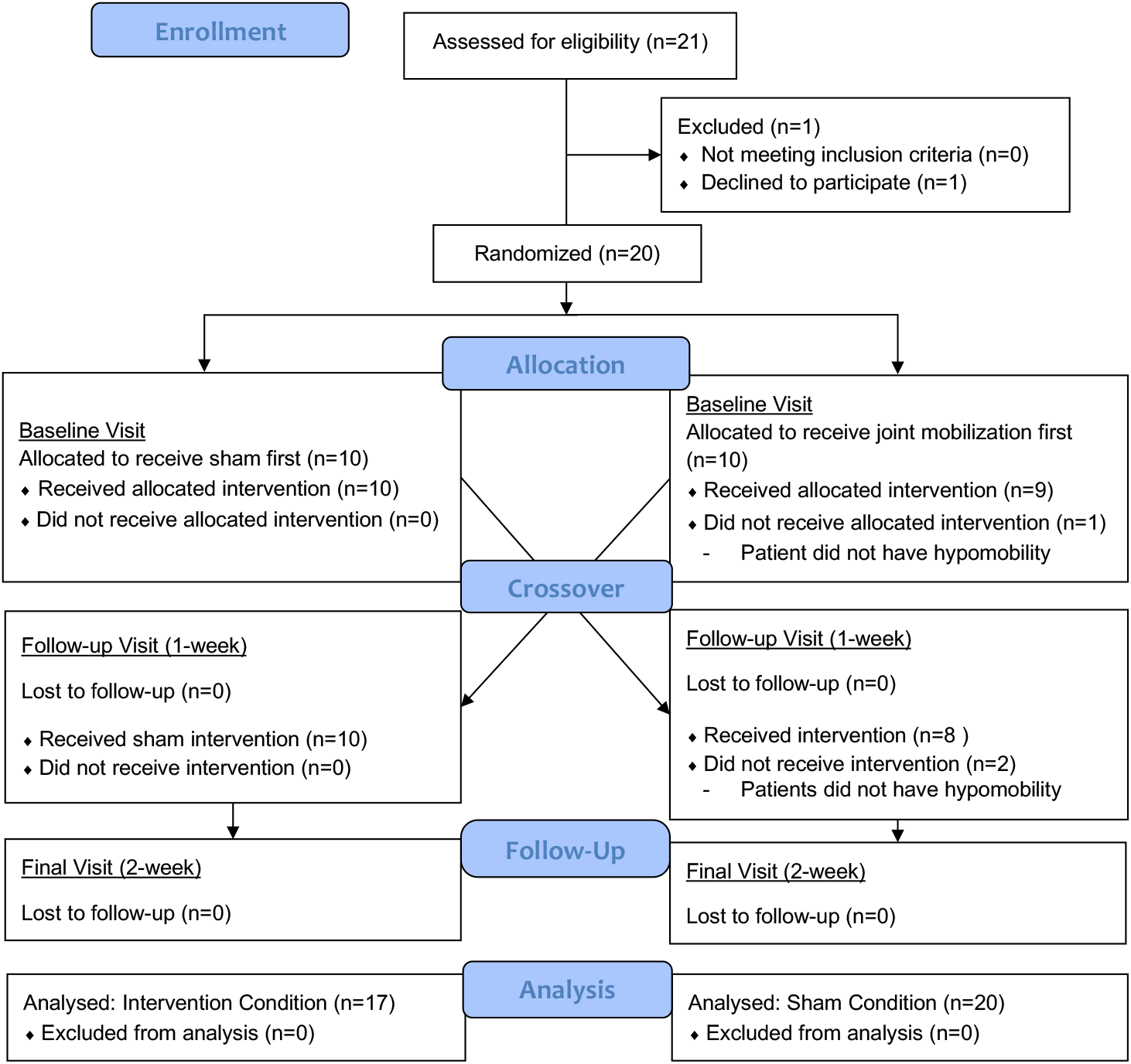
CONSORT Flow Diagram.

### PROCEDURES

#### Baseline Visit

Participants provided demographic information, health and injury history, and completed the FAAM ADL^21^ and Sport subscales,^4^ IdFAI,^5^ the Patient Reported Outcomes Measurement Information System (PROMIS) General Health Questionnaire,^17^ the 11-item Tampa Scale of Kinesiophobia (TSK-11),^27^ and the Godin Leisure-time Exercise Questionnaire.^12^ Height, mass, and true leg length were measured. Foot posture was assessed in standing using the Foot Posture Index–6 item version (FPI), a categorical measure of foot type that is based on five observations and one palpatory assessment.^25^

Demographic, medical history, and FPI assessments were performed by a physical therapist and board-certified orthopaedic clinical specialist with 15-years of clinical experience. Physical examinations were performed by either an athletic trainer with three-years clinical experience or a physical therapist with two-years clinical experience who were blinded to participants’ medical history, functional status, and treatment allocation.

##### Morphologic Foot Assessment

Morphologic foot measurements were obtained using the Arch Height Index Measurement System (JAKTOOL Corporation, Cranberry, NJ). Total and truncated foot length, arch height, and foot width were measured in sitting and standing. Test-retest reliability for these measures were previously reported by the authors to be excellent.^10^ Arch height index^3^ and foot mobility magnitude^22^ were calculated using the component measurements across loading conditions.

##### Joint Motion Measures

Weight bearing dorsiflexion (WBDF)^1^, ankle plantarflexion, inversion, and eversion, and forefoot inversion and eversion joint motion measures^10^ were performed using previously described methods. WBDF was reported as the linear distance measured from the wall to the toes in centimeters. Joint motion measures of rearfoot plantarflexion, inversion, and eversion were performed using a 30.5-cm plastic goniometer (Merck Corporation, Kenilworth, NJ) and reported in degrees. Forefoot inversion and eversion was measured using a digital inclinometer (Fabrication Enterprises, White Plains, NY) and reported in degrees. Linear excursion of first metatarsal (MT) dorsiflexion and plantarflexion were measured utilizing a custom measuring device consisting of two metal rulers bent to 90° and reported in millimeters.^13^ Test-retest reliability for these measures were previously reported by the authors to be good to excellent.^10,13^

##### Muscle Strength

Ankle dorsiflexion, plantarflexion, inversion, eversion, and hallux flexion and lesser toe flexion strength were assessed with a handheld dynamometer (Hoggan Health Industries, West Jordan, UT).^10^ For toe flexion strength measures, the ankle was positioned in 45° plantarflexion to reduce contribution of the extrinsic foot muscles and increase demand of the intrinsic foot muscles.^16^ Strength measures were based on the highest value of three trials. An estimate of torque was derived from the product of force and segment length, normalized to body mass, and reported in Nmkg^-1^. Test-retest reliability for these measures were previously reported by the authors to be excellent.^10^

##### Dynamic Balance

Dynamic balance was assessed using the anterior, posteromedial, and posterolateral directions of the Star Excursion Balance Test (SEBT),^18^ a measure has been found to have excellent test-retest reliability.^20^ Reach distance was normalized to leg length.^15^

#### Intervention

Following baseline assessment, all participants were instructed in a HEP consisting of triceps surae stretching; four-way stretch of the rearfoot, midfoot, and forefoot; isotonic inversion, eversion and dorsiflexion exercises against resistance tubing; single-limb heel raising; and a single limb balance exercise (**Figure 2**).^7,11^ Participants were asked to perform all exercises thrice daily, were provided a handout detailing the exercises, and verbalized understanding following instruction. The decision to utilize a HEP over supervised rehabilitation was to better elucidate the specific treatment effects of the midfoot joint mobilizations.^11^

**Figure 2.** Home exercise program. From Fraser JJ, Hertel J. Preinjury to postinjury disablement and recovery after a lateral ankle sprain: A case report. Journal of Athletic Training. 2018;53(8):776-781. doi:10.4085/1062-6050-114-17. Used with permission of the publisher.

Participants were randomized *a priori* using a random number generator by the senior author and stratified by sex to receive either the midfoot joint mobilizations or sham intervention on the initial visit. Allocation was performed by an otherwise uninvolved laboratory assistant, concealed in a sealed opaque envelope, and opened by the treating clinician who was a board-certified orthopaedic physical therapist with 15-years of clinical experience. Participants allocated to receive midfoot mobilizations were provided a dorsolateral cuboid glide with forefoot supination and 1^st^ tarsometatarsal plantar glides.^6^ Each mobilization technique was an oscillatory Maitland Grade IV applied for 30-seconds duration. If cavitation was not experienced during the first bout of oscillations, a second 30-second bout was provided. In the case where the participant did not exhibit midfoot hypomobility on physical examination (n=3), joint mobilizations were deferred, and the participant was provided the sham treatment only. Participants allocated to the sham treatment were told that they were to receive a gentle soft-tissue technique similar to massage and were provided a “laying of hands” for 30-seconds using the same hand position and contacts used for the joint mobilizations. Participants rated the change of symptoms using a single assessment numeric evaluation (SANE, −100%=full exacerbation, 0=no change, 100%=full resolution) immediately post-intervention and completed the Global Rating of Change (GROC, −7= A very great deal worse, 0= About the same, 7= A very great deal better).

#### Follow-up Visit

Participants returned to the laboratory following a one-week washout for reassessment. They completed the PROMIS, Godin, FAAM-ADL and Sport, SANE, and GROC. HEP compliance was assessed by having the participants demonstrate the instructed exercises. Participants were rated by the treating clinician whether or not they could demonstrate the exercises without hesitation and with appropriate technique. Participants self-rated their compliance using a SANE, with 0% reflective of complete non-compliance with all home exercises and 100% representing performance of all exercises thrice daily. Any deficiencies in exercise technique were corrected and participants were provided encouragement to continue.

Repeat physical examinations were performed pre-and post-intervention. Following the pre-intervention physical examination, participants crossed over to receive the second intervention (i.e. individuals who initially received the sham intervention now received the midfoot joint mobilizations). Participants rated treatment response (SANE) immediately post-intervention and at the end of the visit and completed the GROC.

#### Final Visit

Participants returned to the laboratory one-week later for the final reassessment visit consisting of HEP compliance, patient-reported outcomes, and physical examination.

## STATISTICAL ANALYSIS

*A priori* sample size estimation of 14 participants were needed based on an anticipated 15-point change in the FAAM Sport, an *α*=.05, and *β*=.20.^19^ Descriptive statistics were calculated for demographic and self-reported measures for each subset of the sample allocated to receive either sham or midfoot mobilization during the first visit. Effectiveness of the two interventions (midfoot joint mobilization, sham) and the order of treatments were assessed using multivariate linear regression. Ordinal measures that had greater than five items (GROC) were treated as continuous data during analysis.^24,26^ Participants were analyzed per allocation using intention to treat. Data was analyzed using R Version 3.5.1 (The R Foundation for Statistical Computing, Vienna, Austria). The level of significance was *p* ≤.05 for all analyses.

## RESULTS

Self-reported compliance with the HEP was high following both interventions at Week 1 (Mobilization intervention first: 65.9±25.6%; Sham intervention first: 69.6±22.2%; Hedge’s g: 0.15±0.98) and Week 2 (Mobilization intervention first: 71.9±17.3%; Sham intervention first: 61.3±26.7%; Hedge’s g: 0.45±0.96). When asked to demonstrate the home program, the group that was provided the sham treatment first had a substantially higher proportion of the sample that were able to recall and perform the HEP (75.0%) compared to the participants that initially were provided the midfoot mobilization (33.3%). At Week 2, the ability to recall and perform the HEP was more consistent between groups (Sham intervention first group: 66.7%; Mobilization intervention first group: 83.3%).

Table 2 details the baseline and change measures for patient-reported outcomes of pain, function, and perceived improvement. Participant demonstrated significantly greater perceived improvement immediately posttreatment in the single assessment numeric evaluation (Sham: 5.0±10.2%; Mobilization: 43.9±26.2%; β: 6.8 p<0.001, Adj R^2^:0.17) and Global Rating of Change (Sham: −0.1±1.1; Mobilization: 1.1±3.0; β: 1.8 p=0.01, Adj R^2^:0.12). Tables 3 and 4 details the baseline and change measures for ankle-foot morphological, joint mobility, neuromotor function, and dynamic balance outcome measures following the midfoot mobilization, regardless of order. Additionally, greater improved rearfoot inversion mobility (Sham: 4.4±8.4°; Mobilization: −1.6±6.1°; β: −6.37, p=0.01, Adj R^2^:0.19), plantarflexion mobility (Sham: 2.7°±6.4; Mobilization: −1.7°±4.3; β: −4.36, p=0.02, Adj R^2^:0.07), and posteromedial dynamic balance (Sham: 2.4±5.9%; Mobilization: 6.0±5.4%; β: 3.88, p=0.04, Adj R^2^:0.10) was observed following midfoot mobilization compared to the sham at 1-week post-treatment. Order of interventions was not a significant factor for any of the outcome measures. There were no other significant findings.

**Table 2.**
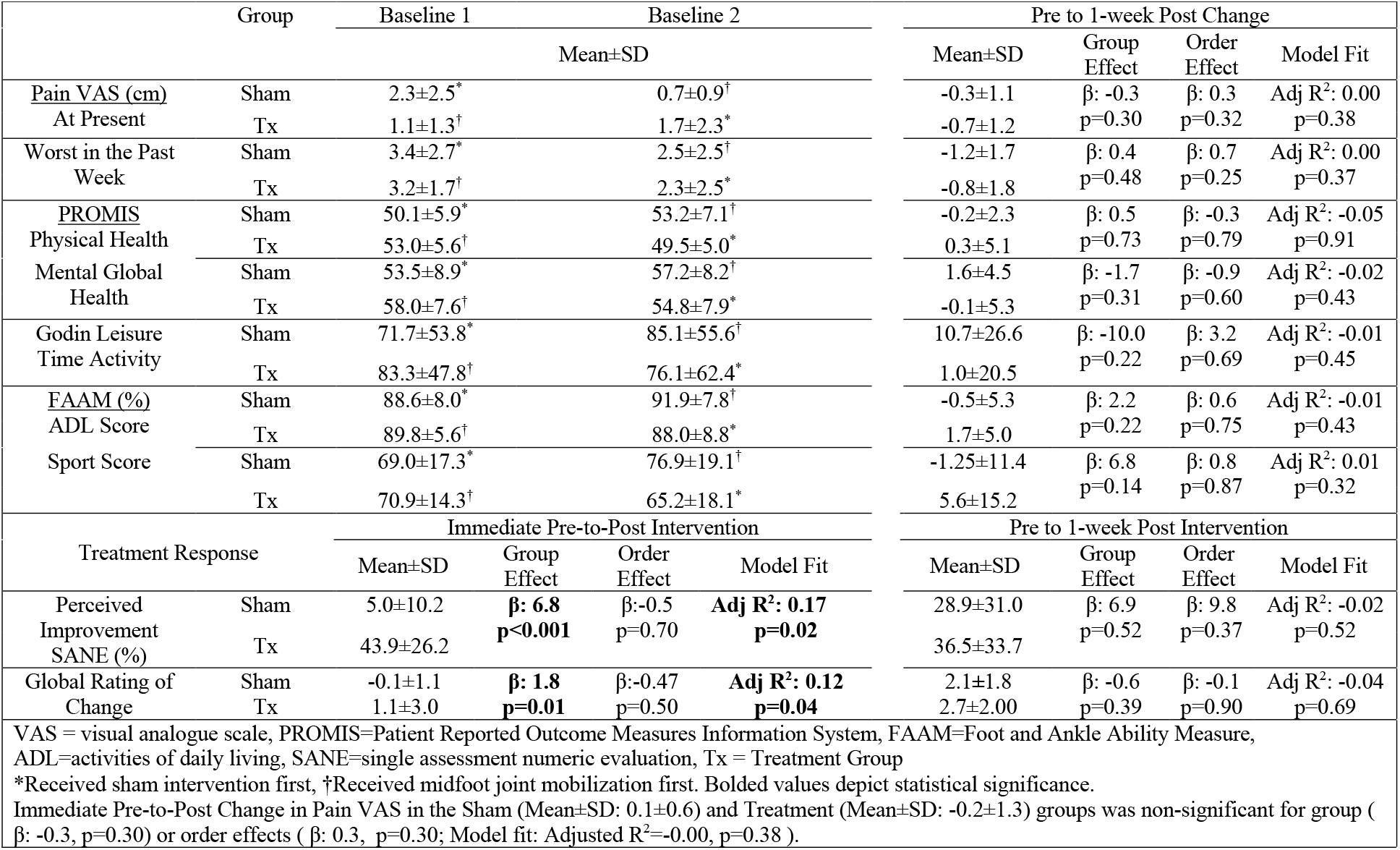
Comparison of treatment on change in patient-reported outcome measures in individuals with chronic ankle instability

**Table 3.**
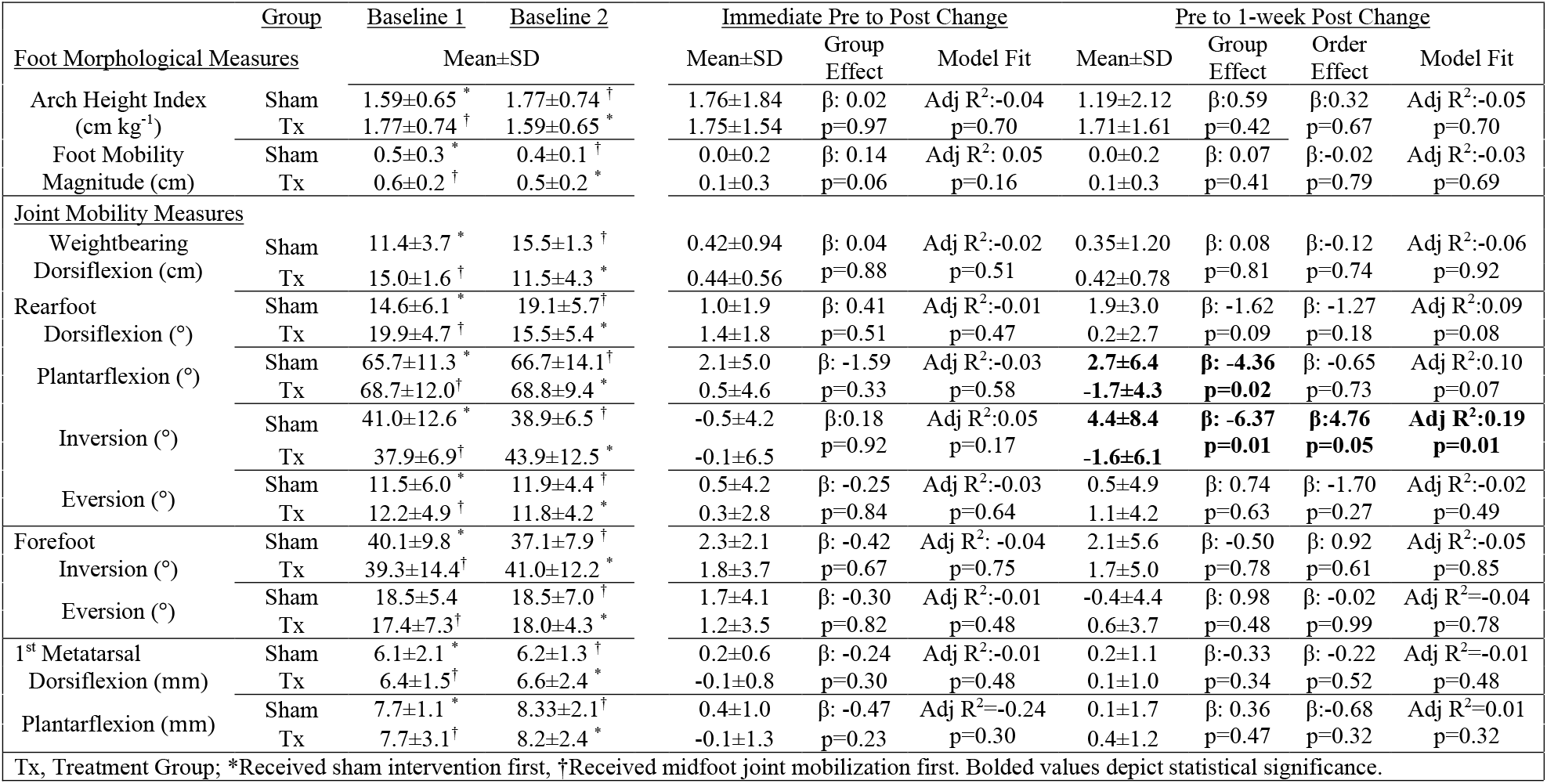
Comparison of treatment on change in ankle-foot joint morphological and mobility outcome measures in individuals with chronic ankle instability

**Table 4.**
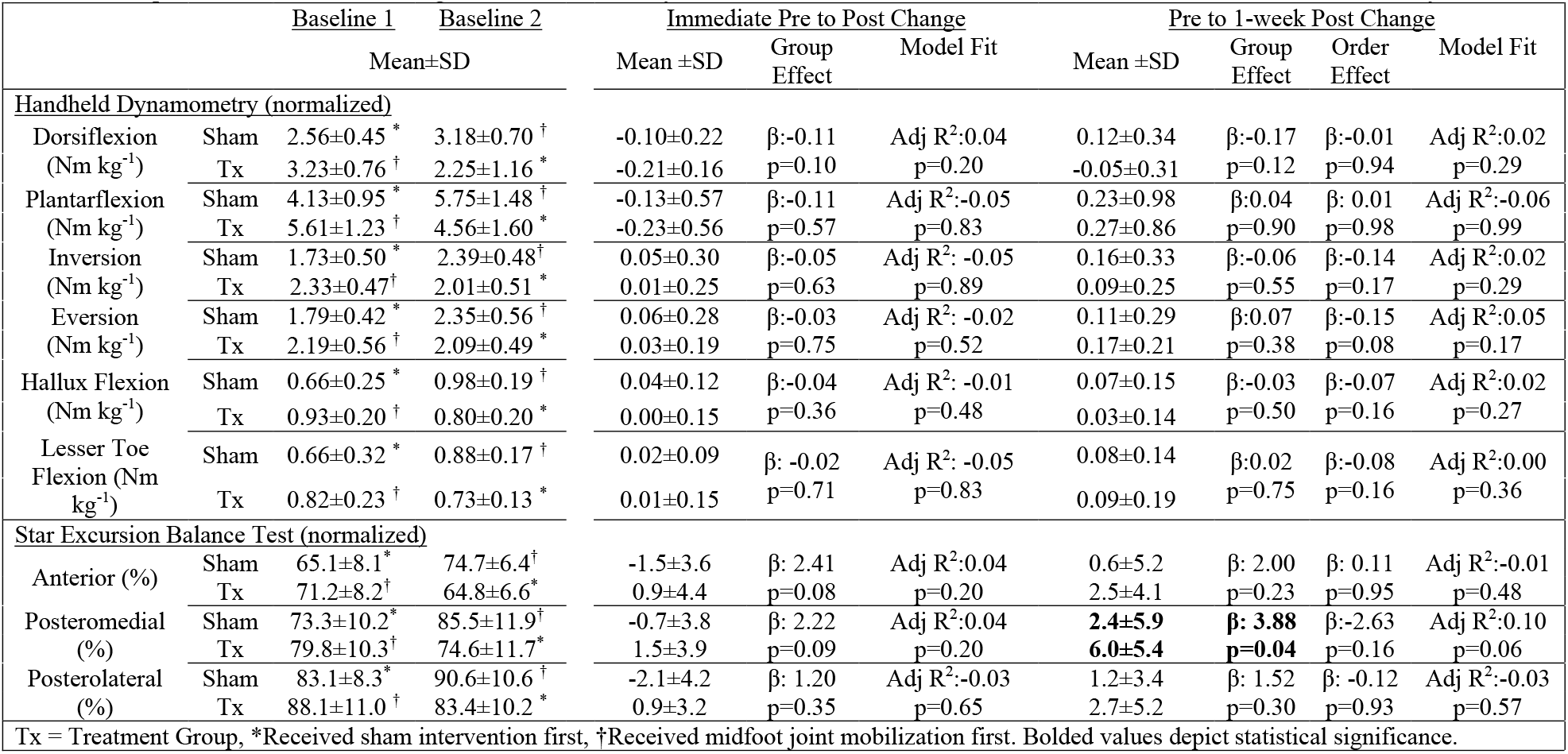
Comparison of treatment on change in neuromotor and dynamic balance outcome measures in individuals with chronic ankle instability

## DISCUSSION

The primary findings of this crossover clinical trial were that CAI patients who received midfoot joint mobilization and a HEP resulted in greater perceived improvement and global rating of change immediately following treatment compared to the receipt of a sham treatment and HEP. At one-week post treatment, patients receiving the midfoot joint mobilization also demonstrated reduced rearfoot inversion and plantarflexion joint mobility and improvements in posteromedial reach distance in the assessment of dynamic balance, whereas those receiving the sham treatment did not.

The findings of greater perceived improvement study are consistent with what was observed in the related study of midfoot mobilization in individuals with subacute LAS. ^11^ MT is purported to work through the interplay of biomechanical, psychological and neurophysiological mechanisms.^12^ In the related study assessing midfoot mobilization in individuals with LAS, the experimental intervention similarly yielded moderate to large magnitude perceived improvements compared to the sham intervention. In the updated model of CAI, psychological status has been highlighted as an important mediator for functional outcomes.^2^ This is substantial since individual with CAI have been found to have increased pain-related fear^13^, a finding that similarly observed in our sample.

While there were only trivial and non-significant improvements in dynamic balance immediately post-treatment, there was a meaningful significant improvement observed one week after receipt of mobilization. Previous study of MT in this population found that rearfoot joint mobilizations and plantar massage increased dynamic balance through likely mechanical and neurophysiological mechanism, improvements that persisted up to one-month following treatment.^14^ Due to the delay in observed improvements, other plausible mechanisms may explain our findings. Specifically, improvements in psychological status may have influenced the changes in dynamic balance, a salient factor that has been found to mediate functional movement performance.^15^ Mitigation of fear in patients with CAI has been associated with improvements in physical performance outcomes, such as dynamic balance.^16^ Furthermore, improved psychological readiness resulting from increased knowledge of testing procedures, beliefs of improvement, and attitudes toward performance execution following the experimental intervention quite possibly influenced these outcomes through improved self-efficacy.^17^

A small, but significant, decrease in rearfoot inversion mobility (mean: −1.6°, 95% CI: −4.4 to 1.2°) and plantarflexion mobility (mean: −1.7°, 95% CI: −3.7 to 0.3°) was observed 1-week following joint mobilization, whereas the sham treatment resulted in an increase in these measures (inversion, mean: 4.4°, 95% CI: 0.7 to 8.1°; plantarflexion, mean: 2.7°, 95% CI: 0.8 to 4.6°). Individuals with CAI have been found to have a more inverted and plantarflexed rearfoot during functional activities such as walking.^18^ It is plausible that reduced rearfoot joint motion may be advantageous and result from improved midfoot mobility and rearfoot mechanics during function in the week following treatment. Restoring normal ankle-foot biomechanics may help to reduce the abnormal stresses placed on the ligaments of the complex and prevent further injuries.^19^ These suppositions require substantiation in future study.

## CLINICAL RECOMMENDATIONS

Higher perceived improvement was observed in the treatment group following joint mobilization compared to sham. This is clinically meaningful because patient preferences form an integral component of evidence-based medicine when it comes to informed clinical decision making.^20^ There are wide range of emotions reported after the injury that may include anxiety, fear, and anger. Some athletes may be affected more by the severity of the injury translating in to depressed mental state. This emotional toll that an athlete goes through because of injury has certain implications in rehabilitation and may persist beyond rehabilitation programs.^21^ It has been suggested that because of an injury, athletes may fall in to a vicious cycle of a general lack of movement that may result in decrease in strength and reinjury which makes them more prone to reinjury. ^21^ While there were only modest physical effects observed with the experimental intervention, the positive psychological effects of the inclusion of midfoot joint mobilization may help to bolster a comprehensive rehabilitation program when treating individuals with CAI. Positive psychological characteristics such as high resiliency and self-efficacy are considered of seminal importance in rehabilitation and return to activity.^22^ Lastly, based on the proportion of CAI patients with observed side to side midfoot hypomobility observed in this study, it is imperative that clinicians examine and treat all the segments of the ankle-foot complex when managing this patient population.

## LIMITATIONS

There are limitations to this study. One of the primary limitations of the cross-over designs is the potential for carryover effects. Comfort maybe taken that we did not observe any significant order effects or treatment by order interactions in any of our primary outcomes. In addition, the choice to use change scores was made *a priori* to mitigate potential carryover effects. Outcomes measures were assessed immediately and one week post mobilization. It is unclear if any observed improvements persisted beyond the study epoch. Therefore, future studies that assess long-term outcomes is warranted. Also, while the use of single mobilization treatment in conjunction with a 1-week HEP was purposeful to specifically assess the perceptual and physical effects of the specific treatment, this does not reflect the standards of practice. As such, external validity of these finds are limited. More research is warranted to investigate the effects of mid-foot mobilization incorporated in a comprehensive rehabilitation program. The time between the application of the intervention and the follow up was deliberately short to mitigate the effects of time and healing. This delimitation precluded assessment of potential longer term effects. Finally, we used a range of outcomes measures to account for potential improvements in physical impairment and activity, in addition to psychological mediators. This could potentially increase the risk of Type I error. Acknowledging this risk, we did not rely solely on p-values for determining treatment effectiveness and also considered the magnitude of change in our interpretation of results.

## CONCLUSION

A single session midfoot joint mobilization, when used in conjuction with a HEP consisting of stretching, strengthening, and balance was highly effective in improving patient’s perceived improvement when compared to a sham treatment. In addition, modest improvements in dynamic balance and rearfoot inversion and plantar flexion motion were also observed in the experimental group. Integration of midfoot joint mobilization should be considered as part of a larger comprehensive rehabilitation program for individuals with CAI.

## Data Availability

All data produced in the present study are available upon reasonable request to the authors.

## Acknowledgements

Stephan Bodkin, PhD, ATC for his assistance with allocation. Navy Medicine Professional Development Center and the University of Virginia’s Curry School of Education Foundation for their generosity in providing funding.

